# SLPI and CXCL10 in non-obese patients, and Serpin E1 in obese patients as AKI biomarkers after cardiac surgery

**DOI:** 10.1101/2025.11.21.25340764

**Authors:** Naomi Brown, Hayan Pan, Nikol Sullo, Bryony Eagle-Hemming, Kristina Tomkova, Lathishia Joel-David, Tracy Kumar, Hardeep Aujla, Alison H Goodall, Gavin J Murphy, Marcin J Woźniak, Bin Yang

## Abstract

**Introduction:** Acute kidney injury (AKI) is a common complication after cardiac surgery, affecting 18% of patients and significantly increasing the risk of mortality. AKI triggers the release of proinflammatory molecules during cardiopulmonary bypass. Obesity, characterised by chronic low-grade inflammation, is an established risk factor for AKI; however, the association between obesity, inflammation and AKI is complex and poorly understood. We investigated the utility of inflammatory molecules in the diagnosis of AKI post-cardiac surgery in obese and non-obese patients.

**Methods:** A panel of 14 circulating plasma molecules (CXCL1, 10 and 13, CCL22, IL-5, 6, 8, 10 and 16, SLPI, TIM1, Properdin, Serpins E1 and A3) was measured using MAGPIX analyser or ELISA kits in samples collected from 95 MaRACAS patients (NCT02315183) before and 6-96 hours after surgery.

**Results:** There was no significant difference in the incidence of AKI between obese (46%) and non-obese (58%) patients. In non-obese patients with AKI, levels of CXCL10, CXCL13, SLPI, and IL-16 were all significantly higher immediately after surgery. SLPI showed the strongest and most consistent association with AKI, with significant predictive value at 6-12, 24 and 72 hours (ROC AUCs 77.7%, 86.3%, and 89.8%). CXCL10 was also associated with AKI at 6-12 hours and 48 hours (ROC AUCs of 84.5% and 72.9%). In obese patients with AKI, Serpin E1 was significantly lower before surgery, but increased at 6-12 hours, showing a significant association with AKI (ROC AUCs 66.7% pre-op and 71.2% at 6–12 hours). Properdin was also significantly higher in obese patients with AKI at 72 hours (ROC AUC of 85%). Other measured biomarkers did not differ between AKI and non-AKI groups in either obese or non-obese patients.

**Conclusions:** Distinct dynamic changes of SLPI and CXCL10 in non-obese and Serpin E1 and Properdin in obese patients have diagnostic potential as biomarkers of AKI post-cardiac surgery. These proinflammatory molecules should be further validated in larger cohorts.

## Introduction

Acute kidney injury (AKI) is a common complication of cardiac surgery that occurs in up to 30% of patients. ^1^ Patients who develop AKI following cardiac surgery suffer from increased morbidity and mortality, prolonged hospital stays and increased healthcare expenditure. ^2,3^ AKI is defined by a sudden decline in renal function, marked by a significant decrease in glomerular filtration rate (GFR). Clinical monitoring of AKI involves measuring urine output and serum creatinine (SCr) levels. However, SCr limits timely diagnosis and lacks prognostic sensitivity and accuracy. ^4^ This necessitates an investigation into novel biomarkers that facilitate earlier AKI diagnosis and intervention.

The pathophysiological mechanisms underlying AKI are complex and poorly understood. It has been reported that cardiopulmonary bypass during cardiac surgery can trigger the release of damage-associated molecular patterns (DAMPs) that activate proinflammatory pathways and further contribute to renal damage. ^5^ This systemic inflammatory response involves the release of cytokines and chemokines, leading to leukocyte infiltration, tubular cell injury, and ultimately inducing AKI. ^5,6^ Whilst various inflammatory biomarkers, including different interleukins, Neutrophil Gelatinase-Associated Lipocalin (NGAL), and T-cell immunoglobulin and mucin-domain containing-1 (TIM1), have been found to predict and diagnose AKI after cardiac surgery ^7–9^, our previous research identified no significant differences in circulating inflammatory biomarkers between AKI and non-AKI cardiac surgery patients. ^10^ Alternatively, our study revealed elevated platelet activation and platelet-leukocyte aggregation in AKI patients ^10^, suggesting a potential role for platelet-mediated mechanisms in the development of AKI after cardiac surgery.

In addition, obesity is a risk factor for various adverse health outcomes, including AKI. ^11,12^ Patients with obesity exhibit chronic low-grade inflammation, which is characterised by elevated levels of inflammatory biomarkers. ^7,13^ This chronic inflammatory state can contribute to endothelial dysfunction, impaired renal autoregulation, and alter the inflammatory response to surgical stress. ^11,14^ Given the established role of inflammation in AKI pathophysiology, we looked to further investigate the differential responses of proinflammatory biomarkers in an AKI population stratified by obesity. ^7,8,10^

Although various inflammatory molecules have been identified as AKI biomarkers, their ability to predict AKI remains limited. ^15^ Secretory leukocyte protease inhibitor (SLPI) is an anti-protease and alarm inflammatory protein that has been found to be elevated in both obesity and AKI, respectively. ^16–18^ SLPI has also been reported as a potential post-operative biomarker of AKI after thoracoabdominal aortic aneurysm repair. ^17^ Alternatively, serpin E1 is a key protein associated with obesity and is notably overexpressed in patients with AKI. ^19,20^ However, no prospective studies have evaluated circulating levels of SLPI or Serpin E1 in AKI patient populations stratified by obesity.

We hypothesised that the obesity-specific proinflammatory molecules, SLPI and Serpin E1, would be upregulated in obese AKI patients and would serve as potential biomarkers of post-cardiac surgery AKI. To test this hypothesis, we conducted a longitudinal analysis of 14 circulating plasma molecules in the plasma of obese and non-obese AKI patients undergoing cardiac surgery.

## Methods

### Study design and setting

An Observational Case Control Study to Identify the Role of MV and MV Derived Micro-RNA in Post CArdiac Surgery AKI (MaRACAS) was a prospective observational study registered at https://clinicaltrials.gov/ct2/show/NCT02315183. The study was approved by East Midlands – Leicester South Research Ethics Committee (REC reference 13/EM/0383) and ran between 11th December 2014 and 30th May 2017 at Glenfield General Hospital, Leicester, UK.

### Study Cohort

The study cohort included 95 adult (>16 years) cardiac surgery patients undergoing coronary artery bypass grafting (CABG) or open-valve surgery. All participants were at an increased risk for AKI, determined by a modified AKI risk score. ^21^ The exclusion criteria were as follows: emergency or salvage procedure, critical pre-operative state (Kidney Disease: Improving Global Outcomes (KDIGO) Stage 3 AKI ^22^, requiring ventilation, ionotropes, or intra-aortic balloon pump), pre-existing inflammatory state (sepsis undergoing treatment, chronic inflammatory disease, AKI within 5 days or congestive heart failure), ejection fraction <30%, or pregnancy. Obesity was defined as BMI≥32 based on our previous analysis of gene expression profiles. ^23^

### Sampling and storage

Blood samples were collected from patients pre-operatively, upon return to the intensive care unit, and at 6-12, 24, 48, 72, and 96 hours post-operatively. The total blood volume collected was 104.8 ml in clotting, EDTA, citrate and hirudin tubes (S-Monovette, Sarstedt, Neumbrecht, Germany). Plasma samples were collected and stored at -80 °C within 2 hours of collection. Urine samples were collected pre-operatively and 24 hours after surgery. Routine serum creatinine measurements were conducted in NHS laboratories.

### Reducing bias

Laboratory staff were blinded to patient samples that were identified only by trial acronym, patient study ID and time of sample collection. NHS laboratory personnel conducting routine measurements were unaware that samples were collected for a trial. The AKI status of participants was blinded to all researchers during further analysis.

### Soluble biomarker analysis

Levels of plasma biomarkers, CXCL1, 10 and 13, CCL22, interleukin (IL)-5, 6, 8, 10 and 16, secretory leukocyte peptidase inhibitor (SLPI), T cell immunoglobulin mucin domain-1 (TIM1), Properdin and Serpin E1 were measured by multiplex assay on the MAGPIX Analyser (Luminex, Netherlands). Serpin A3 was measured using the human Alpha 1-Anti-Chymotrypsin ELISA Kit (Abnova, Taiwan). Urinary NGAL was measured using the human NGAL ELISA Kit (EKF Diagnostics, UK).

### Statistical Analysis

Data were analysed using R Studio 4.4.1. and plots were prepared with the ggplot2 R package. ^24^ Comparisons were made between patients with and without AKI or obesity. The distribution of baseline variables was determined using the Shapiro-Wilk test. Normally distributed variables are reported as mean ± standard deviation (SD), and non-normally distributed variables are reported as median (interquartile range, IQR). The Chi-Squared (χ2) test was used to assess associations between categorical variables. Continuous parametric comparisons were performed using Student’s T-test. Non-parametric comparisons were performed using the Wilcoxon rank-sum test. All soluble biomarkers were log-transformed (log pg/mL) for comparative analysis and adjusted for multiple comparisons using the Benjamini-Hochberg method. Multivariable logistic regression models were used to assess the association of biomarkers with AKI and were fitted with the glm function. Hosmer–Lemeshow goodness-of-fit test was performed with hoslem.test from the ResourceSelection R package (Sólymos and Lele, 2016). ^25^ The Area Under the Receiver Operating Characteristic curves (AUC-ROC) were calculated using the pROC package in R.

## Results

### Participants

Out of 230 cardiac surgery patients screened, 150 were eligible for the study. Fifty-three participants declined consent, and two were excluded after giving consent. A total of 95 patients completed the study, as described previously ^10^; however, the present study stratified patients by obesity status (**Figure 1A**). Missing data for creatinine, urinary NGAL and plasma biomarker measures were 4.1%, 7.4% and 1.3%, respectively.

**Figure 1.**
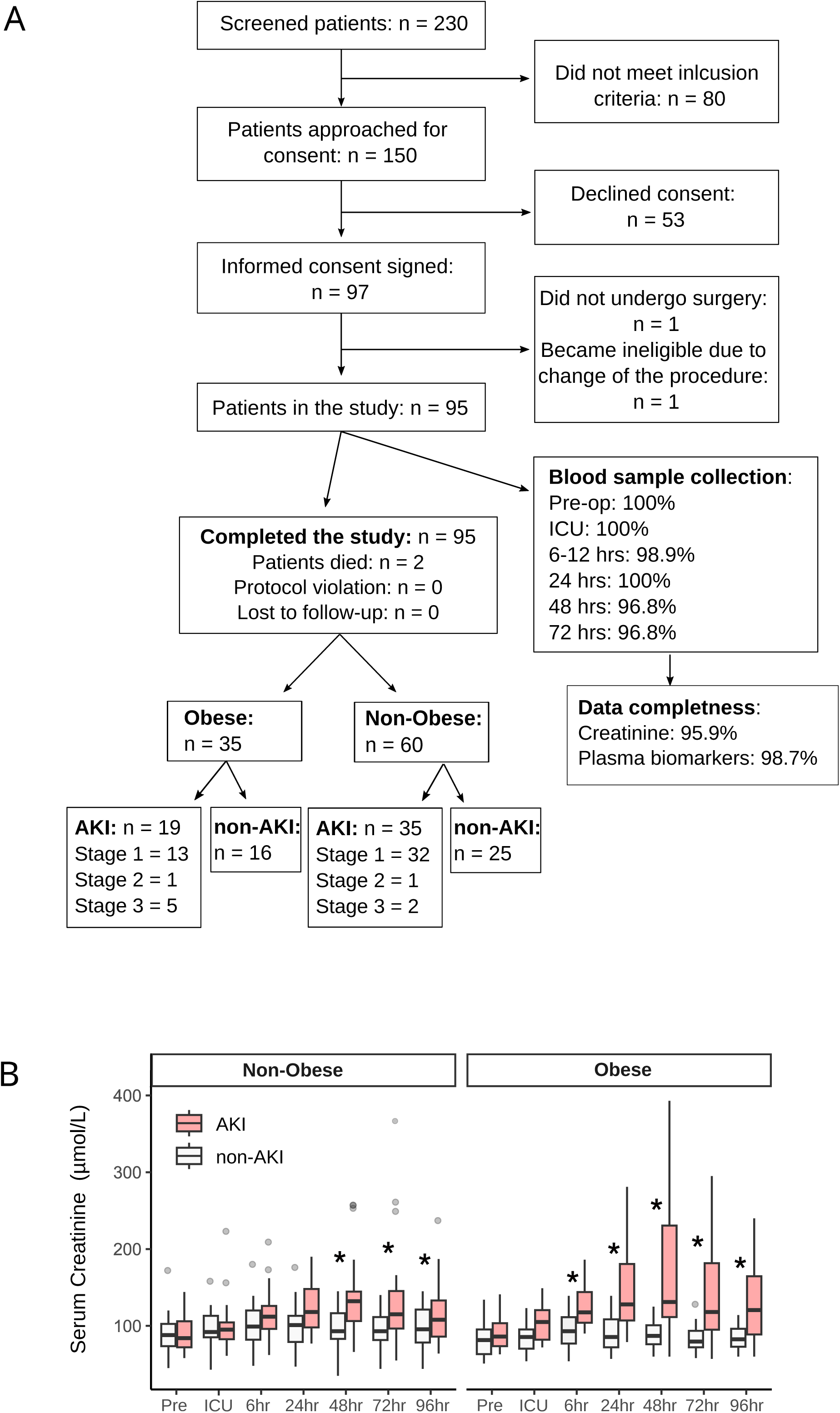
(**A**) CONSORT diagram. (**B**) Levels of serum creatinine. Asterisks indicate a significant difference between no-AKI and AKI (*p*<0.05).

Thirty-five of these patients (37%) were obese, and 60 (63%) were non-obese. AKI was determined according to the KDIGO definition. ^26^ There was no significant difference in the incidence of AKI between obese (19 patients with AKI, 54%) and non-obese patients (35 patients with AKI, 58%) (χ2=0.029, df=1, *p*-value=0.865). The majority of patients exhibited Stage 1 AKI in both the obese (13 patients with stage 1, 68% of AKI patients) and non-obese (32 patients with stage 1, 91% of AKI patients) groups. There was no significant difference in the AKI stage between obese and non-obese groups (χ2=4.748, df=2, *p*-value=0.093). Levels of missing data were less than 5% for the circulating AKI biomarkers (**Figure 1A**).

The baseline demographics of non-obese and obese patients were well matched between the AKI and non-AKI groups. Non-obese patients with AKI were older (75.8 [6.0] vs 71.5 [6.8], *p*-value=0.014) and had longer ventilation times (13 [10-17] vs 9 [8-14], *p*-value=0.01) compared with non-obese non-AKI patients. Obese patients with AKI had significantly longer ventilation time (14 hr [10-21.5] vs 9 hr [7-16.25], *p*-value=0.019), ICU stay (2 days [2-7.5] vs 1 day [1-2.5], *p*-value=0.028) and hospital length of stay (11 days [7.5-15.5] vs 7 days [6-9.25], *p*-value=0.025; **Table 1**) compared with obese non-AKI patients. In addition, the CBP duration was longer in patients with AKI compared with non-AKI for both non-obese and obese groups; however, this was not statistically significant.

**Table 1.**
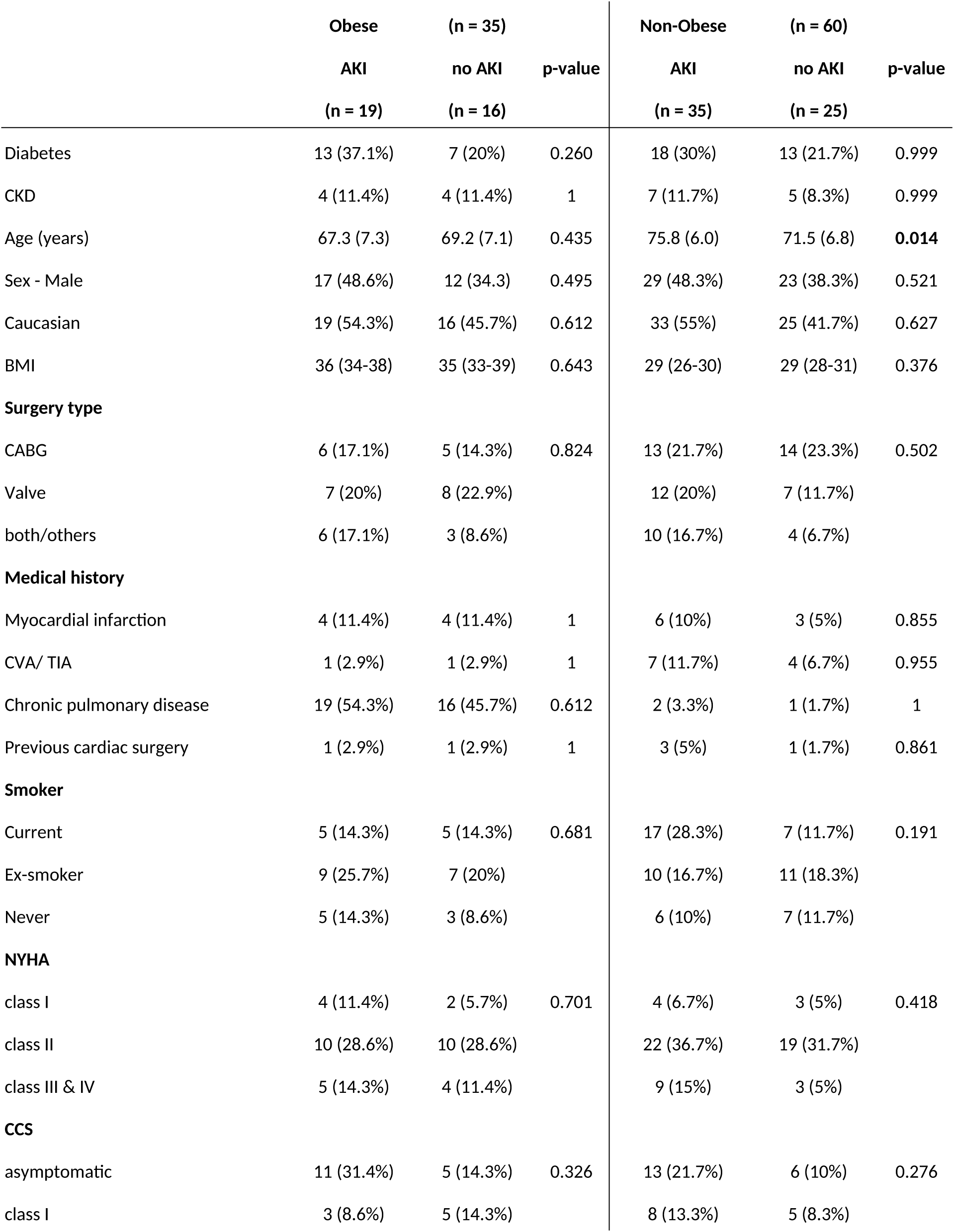
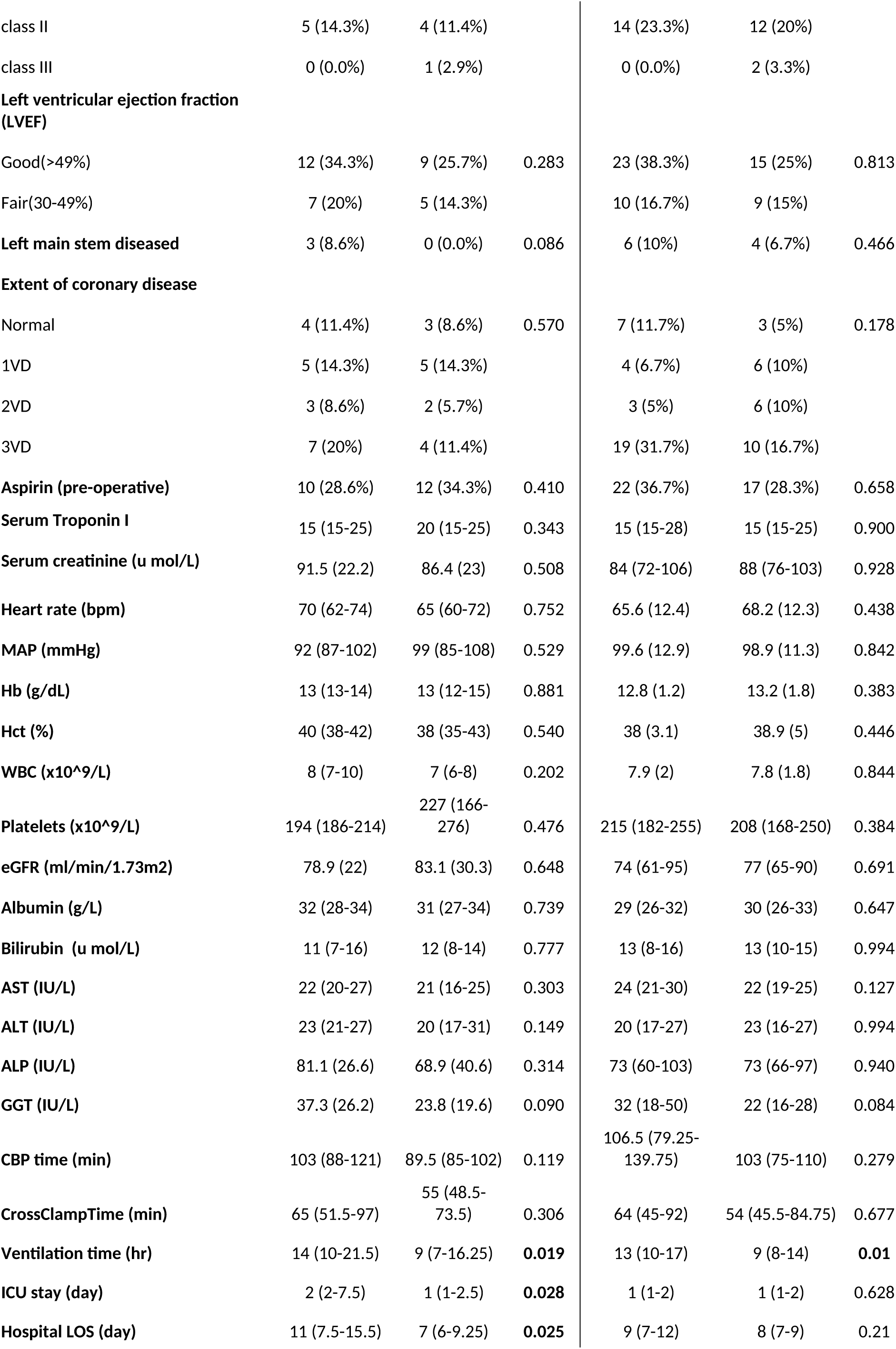
Cohort characteristics. Continuous variables were analysed with t-test or non-parametric Kruskal–Wallis tests. Categorical variables were analysed with exact test. Data are shown as n (%) for categorical variables and mean (standard deviation, STD) or median (interquartile range) for continuous variables. Significant values are given in bold red. * missing data: smoking staus (n=3), CCS (n=1), LVEF (n=5), left main steam diseased (n=4), Extent of coronary disease (n=4)

Levels of serum creatinine peaked 48 hours after surgery for both non-obese (132 [106.3–144.5] µmol/L) and obese (131 [111.5–230.5] µmol/L) AKI patients. In non-obese AKI patients, serum creatinine was significantly elevated 48-96 hours after surgery, whereas in obese AKI patients, elevations occurred earlier and reached higher levels between 6 and 96 hours after surgery (**Figure 1B**). However, there was no difference between AKI and non-AKI groups for the urinary biomarker NGAL in non-obese and obese patients (**eTable 1**).

### Inflammatory biomarkers

Post-operative measurements of several of the inflammatory biomarkers, including CCL22, CXCL1, IL-5, 6, 8 and 10, Serpin A3 and TIM1, showed no significant differences between AKI groups in patients with or without obesity. Subsequent analysis focused on biomarkers that demonstrated clear temporal changes after cardiac surgery.

In the non-obese group, out of fourteen biomarkers, CXCL10, CXCL13, IL-16, and SPLI were significantly higher in patients with AKI. None of these markers were significantly different between AKI and non-AKI patients in the obese group. However, Serpin E1 was significantly lower in patients with AKI before surgery and higher at 6-12 hours afterwards. Properdin levels were significantly lower at 72 hours in obese patients with AKI (biomarker summary in **Figure 2A** and level plots in **Figure 2B**).

**Figure 2.**
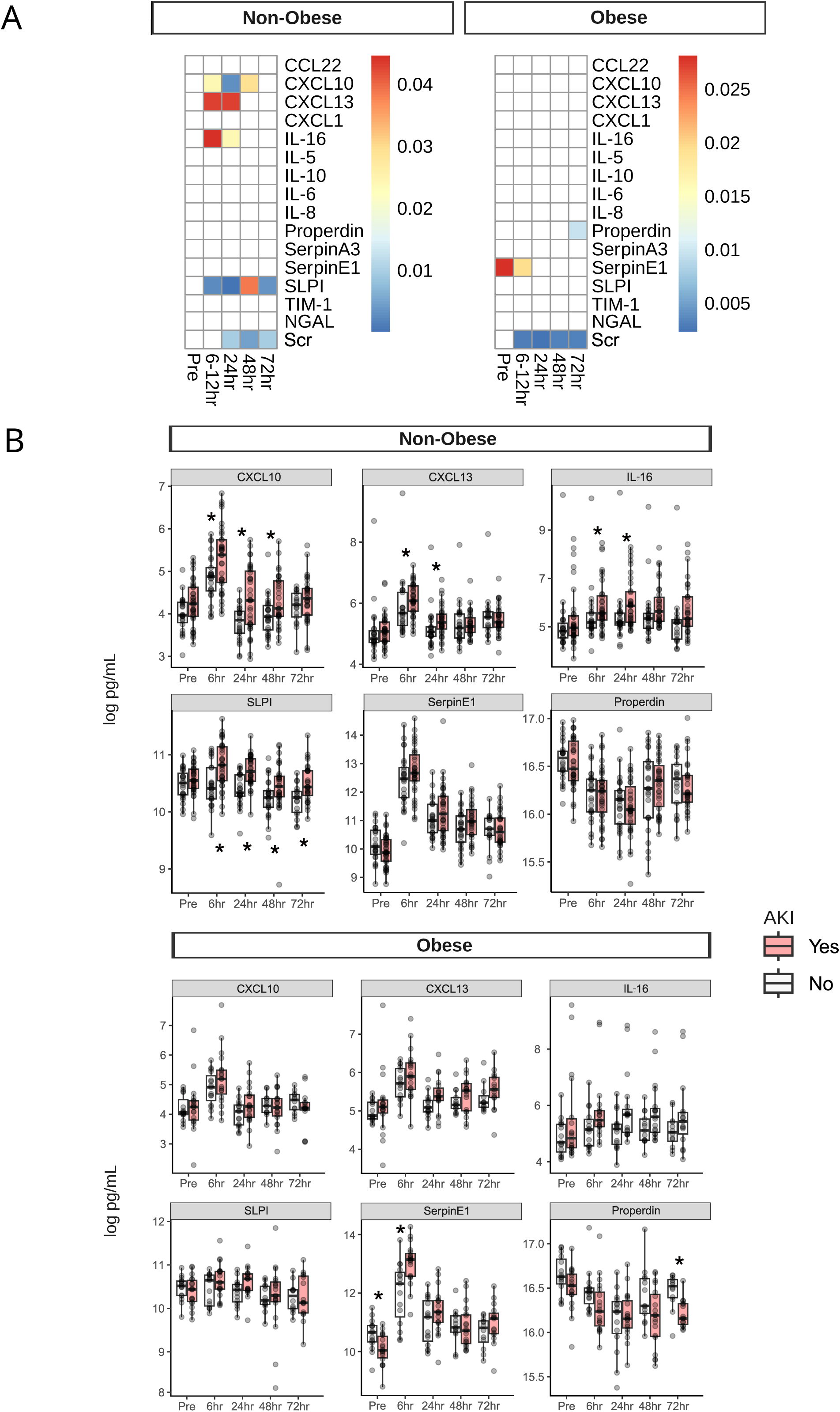
(**A**) Summary of the biomarker analysis (**eTable 1**). The colour codes indicate *p*-value. (**B**) Boxplots of significant variables in non-obese and obese groups. Asterisks indicate a significant difference between no-AKI and AKI (*p*<0.05).

In non-obese patients with AKI, biomarkers CXCL10, CXCL13, interleukin-16 and SLPI changed significantly in contrast to those in the non-AKI group at different time points after surgery. Levels of CXCL10 were significantly higher in the AKI group at 6-12 hours (5.3 ± 0.8 vs 4.8 ± 0.5 log pg/mL, p = 0.024), 24 hours (4.3 ± 0.8 vs 3.8 ± 0.4 log pg/mL, p = 0.003) and 48 hours (4.1 [3.9–4.8] vs 3.9 [3.6–4.2] log pg/mL, p = 0.029), peaking at 6 hours. CXCL13 levels were higher 6-12 hours (6.1 [5.7–6.6] vs 5.7 [5.3–6.4] log pg/mL, p = 0.043) and 24 hours (5.4 [5.2–5.6] vs 5.1 [4.9–5.2] log pg/mL, p = 0.043). Interleukin-16 levels were significantly higher at 6-12 hours (5.5 [5.3–6.3] vs 5.2 [4.9–5.6] log pg/mL, p = 0.045) and 24 hours (5.8 [5.2–6.4] vs 5.2 [5.1–5.6] log pg/mL, p = 0.024). Finally, SLPI levels were higher at 6-12 hours (10.8 ± 0.4 vs 10.5 ± 0.4 log pg/mL, p = 0.003) 24 hours (10.7 ± 0.3 vs 10.4 ± 0.3 log pg/mL, p = 0.0001), 48 hours (10.5 [10.3–10.6] vs 10.3 [10.1–10.4] pg/mL, p = 0.039) and 72 hours (10.5 ± 0.3 vs 10.3 ± 0.3 log pg/mL, p = 0.003), peaking at 6-12 hours.

In the obese AKI patients, levels of Serpin E1 were significantly lower before surgery (10 ± 0.5 vs 10.6 ± 0.5 log pg/mL, p = 0.028) and higher 6 hours (13 ± 0.7 vs 12 ± 1.1 log pg/mL, p = 0.02) post-surgery. In contrast to the other biomarkers, properdin levels were lower in AKI patients at all time points and were significantly lower at 72 hours post-surgery (16.2 ± 0.2 vs 16.5 ± 0.2 log pg/mL, 0.01; **Figure 2B, eTable 1**).

### Diagnostic utility of the inflammatory biomarkers

To gain an insight into the association between the significant variables and AKI in non-obese and obese groups, we fitted multivariable logistic models with a single binary outcome (AKI) with levels of inflammatory biomarkers as predictors. Each model included AKI confounders (age, diabetes and baseline kidney function). The discriminatory ability of significant variables from each model was further tested by fitting ROC curves.

#### Non-obese group

At 6-12 hours, the levels of CXCL10, CXCL13, IL-16 and SLPI were significantly higher in AKI patients (**Figure 2A**). Since all markers significantly correlated with each other, we fitted a logistic model of each of them together with AKI confounders. Only SPLI was significantly associated with AKI (Odds Ratio (OR): 23.58, 95% Confidence Intervals (CI): 3.1-179.07, *p*-value<0.01). In the model, the age of patients was also significant (**eTable 2).** SLPI and age offered fair discriminative ability between AKI and non-AKI (AUC: 77.7%, 65.4%−90.0%; **Figure 3A**).

**Figure 3.**
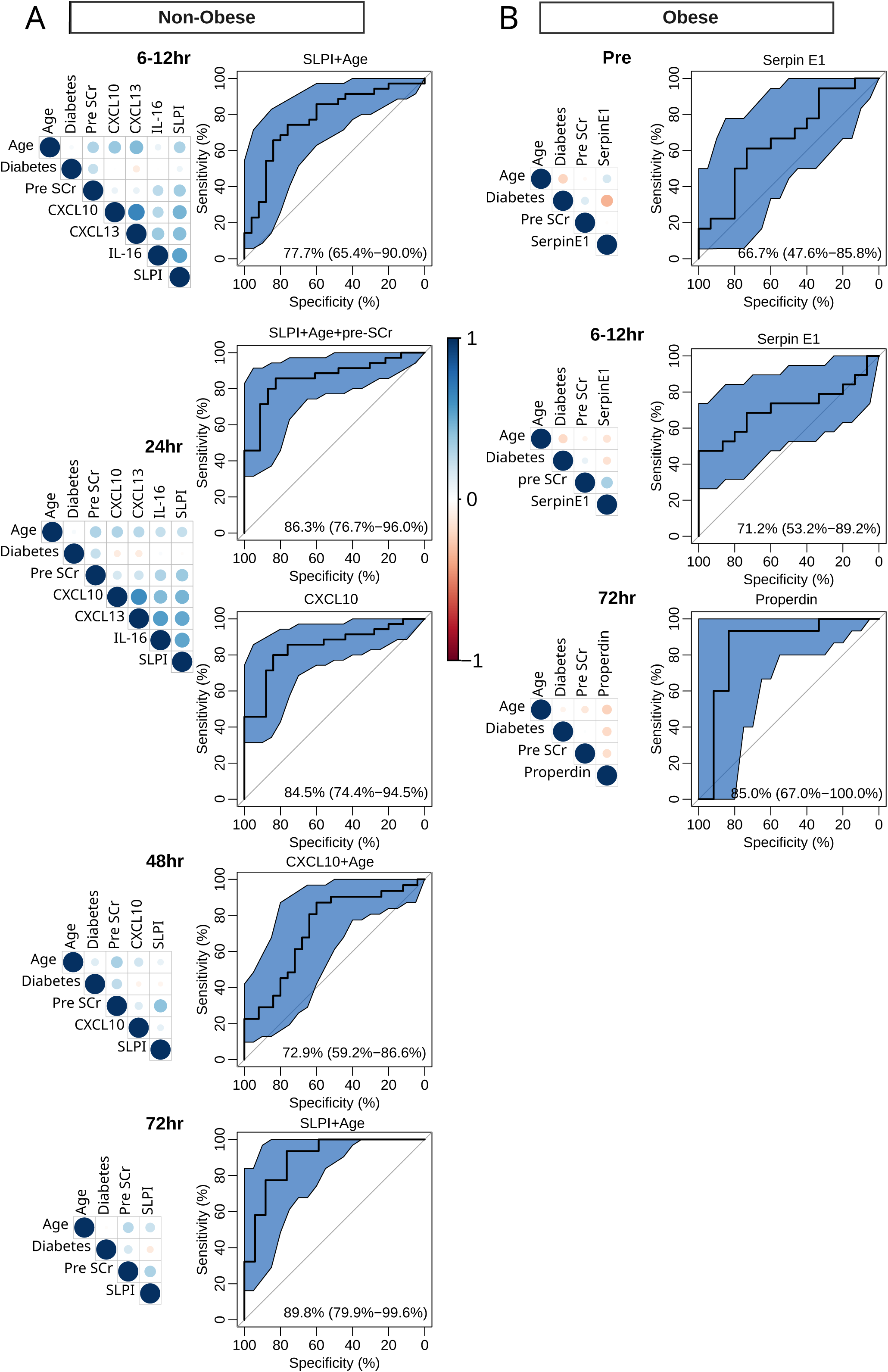
Correlations between significant variables between AKI and non-AKI at each time point in non-obese (**A**) and obese (**B**) groups.

At 24 hours, CXCL10, CXCL13, IL-16 and SLPI were also significantly higher in the AKI group (**Figure 2A**) and significantly correlated with each other. SLPI (OR: 502.07, CI: 15.66-16093.71, *p*-value<0.01) and CXCL10 (OR: 3.91, CI: 1.32-11.55, *p*-value=0.01) were significantly associated with AKI. In the SLPI model, age and pre-operative levels of serum creatinine (preSCr) were also significant (**eTable 2**). Both SLPI with age and preSCr (AUC: 86.3%, 76.7%−96.0%) and CXCL10 (AUC: 84.5%, 74.4%−94.5%) offered considerable discriminative ability between AKI and non-AKI (**Figure 3A)**.

At 48 hours, SLPI and CXCL10 were significantly higher in the AKI patients (**Figure 2A**). Both markers correlated significantly and the models were fitted separately. CXCL10 was significantly associated with AKI (OR: 3.29, CI: 1.01-10.75, *p*-value<0.05) in a model where age was also significant (**eTable 2**). The marker offered fair discriminative ability between AKI and non-AKI (**Figure 3A).**

At 72 hours, only SLPI was significantly higher in the AKI patients (**Figure 2A**). Its levels are significantly associated with AKI (OR: 500.65, CI: 3.16-79200.63, *p*-value=0.02). In the model, patients’ age was also significant (**eTable 2**). The marker offered considerable discriminative ability between AKI and non-AKI (AUC: 89.8%, 79.9%−99.6%; **Figure 3A**).

#### Obese group

Levels of Serpin E1 were significantly lower in AKI patients before surgery and higher at 6-12 hours (**Figure 2A**). The biomarker was significantly associated with AKI at both time points (Pre OR: 0.17, 0.03-1, *p*-value<0.05; 6-12hr OR: 6.81, 1.68-27.61, *p*-value=0.01; **eTable 2**). Serpin E1 offered poor discriminative ability between AKI and non-AK before surgery (AUC: 66.7%, 47.6%−85.8%) and fair at 6-12 hours (AUC: 71.2%, 53.2%−89.2%; **Figure 3B**).

Properdin was significantly lower in AKI patients with AKI at 72 hours (**Figure 2A**), and it was significantly associated with AKI (OR: 1.6×10⁶, CI: 0-0.92; **eTable 2**) and offered considerable discriminative ability for AKI (AUC: 85.0%,67.0%−100.0%; **Figure 3B**).

In addition, the changes of other detected molecules, including TIM1, NGAL and Serpin A3, did not reach statistical significance between the compared groups and time points.

## Discussion

Our study demonstrates that the circulating inflammatory response in cardiac surgery–associated AKI differs between obese and non-obese patients, with distinct biomarker profiles. Out of four biomarkers (SLPI, CXCL10, IL-16, and CXCL13), whose levels were significantly different in AKI, SLPI and CXCL10 significantly associated with AKI in non-obese patients in logistic models. Levels of Serpin E1 (PAI-1) and Properdin were significantly different in obese patients and were also significantly associated with AKI.

### Clinical significance

In non-obese patients, AKI was associated with a rise in proinflammatory markers. CXCL10 (IP-10), a chemokine induced by interferon-γ, increased significantly within 6–48 hours post-surgery in non-obese AKI and was a predictor at 24 and 48 hours. Previous reports identified CXCL10 as a promising injury biomarker in AKI diagnostics. ^27^ SLPI, an anti-protease and anti-inflammatory protein, was also sharply elevated in non-obese AKI (peaking at 6-12 hours) and showed the strongest association with AKI at 6-12 hours. SLPI has renoprotective properties; it mitigates neutrophil protease injury and dampens NF-κB-mediated inflammation. Exogenous SLPI can reduce tubular necrosis, inflammatory cell infiltration, and apoptosis in AKI models. ^28^

Our findings corroborate a prior clinical study that identified SLPI as a novel early biomarker of AKI after cardiac surgery. ^17^ The rise of SLPI in non-obese AKI may reflect a compensatory, nephroprotective response to acute inflammation. SLPI has also been shown to directly protect renal epithelial cells and promote their regeneration after injury. ^28^

CXCL13 and IL-16 also increased in non-obese AKI, suggesting activation of B-cell chemotaxis and T-cell cytokine pathways. CXCL13 transiently increased in ischemia-reperfusion models 24 hours after injury, ^29^ and increases in IL-16 levels were previously associated with renal replacement therapy and death. ^20^ Taken together, the non-obese AKI profile is characterised by an inflammatory burst alongside an upregulation of SLPI that likely reflects an endogenous attempt to counteract inflammation-induced damage.

In obese patients, pre-operative Serpin E1 levels were lower in those who developed AKI, whereas 6-12 hours post-operative Serpin E1 was significantly higher. Serpin E1, also known as PAI-1, is an adipose-derived factor. Obesity is associated with chronically elevated Serpin E1 that can promote a pro-thrombotic, proinflammatory state. The lower pre-operative Serpin E1 in obese patients who developed AKI is counterintuitive and requires further investigation. Serpin E1 is strongly linked to AKI in critical illness, and elevated plasma PAI-1 correlates with AKI risk and severity. ^30^ It can play a dual role, high levels associate with worse outcomes; however, aged mice lacking Serpin E1 suffer more severe AKI and mortality in sepsis, indicating it may aid in injury resolution under certain conditions. ^30^ Therefore, in obese patients, an acute spike in Serpin E1 may reflect an intense insult or a maladaptive response.

Properdin (a positive regulator of complement) did not differ between AKI and non-AKI early after surgery, but by 72 hours post-operative, its plasma levels were significantly lower in obese AKI patients, and it was associated with AKI in logistic models. This can suggest that AKI is accompanied by complement consumption or dysregulation over time. Early post-operative complement activation is expected in all cardiac surgery patients due to cardiopulmonary bypass, potentially masking differences between groups at 6–48 hours. By 72 hours, however, patients with AKI had a significant decline in Properdin levels. Low Properdin has been observed in other inflammatory states (e.g. sepsis non-survivors) and is thought to reflect ongoing complement activation and poorer prognosis. ^31^ Alternatively, Properdin may have a protective role in later-stage AKI repair: in a murine ischemia-reperfusion model, properdin deficiency worsened kidney injury at 72 hours by impairing clearance of apoptotic cells. ^9^ The 72-hours properdin drop in obese AKI patients might indicate a failure to resolve complement-mediated inflammation, aligning with worse renal recovery. The obese AKI profile points to different mediators with comparatively blunted early chemokine/cytokine spikes. Chronic low-grade inflammation in obesity could predispose to these differences. Obese patients have elevated baseline inflammatory markers and may exhibit immune priming or tolerance that alters acute response patterns. ^23^

SLPI and CXCL10, which are predictive of AKI in non-obese patients, point toward acute sterile inflammation as a central driver, involving neutrophil proteases, innate immune activation, and interferon pathways. That SLPI rose in AKI and was protective in experimental AKI ^28^ suggests that non-obese patients are able to raise an anti-inflammatory response, rising a question whether augmenting anti-inflammatory mediators could ameliorate injury. In obese patients, the prominence of Serpin E1 and Properdin shifts attention to coagulation–inflammation crosstalk and the complement system. Increases in Serpin E1 levels might contribute to microvascular thrombosis in the kidney or serve as a marker of endothelial stress. We recently demonstrated that thrombosis plays a significant role in post-surgery AKI. ^10^

The late drop of Properdin in AKI indicates a prolonged complement activation, while a deficiency of Properdin could exacerbate complement dysregulation or impair the clearance of injury debris. ^9^ Although highly speculative, therapies modulating complement (e.g. C5a inhibitors) or Serpin E1 could have different effects in obese and non-obese AKI, and measuring these markers could identify patients with ongoing pathogenic complement activation, which may be potential biomarkers for the prognosis or targeted intervention of AKI.

### Strengths and limitations

The biggest limitation of this study is the sample size (95 patients), which is relatively small, especially after stratifying by obesity. It can limit generalizability and the power to detect smaller effects. Our cohort was derived from a single centre and enriched for high AKI risk, so results may not apply to all cardiac surgery populations. Second, we relied on plasma biomarkers of inflammation; these systemic markers may not be specific to kidney injury and could be confounded by extracorporeal circulation, surgical trauma, or other organ injury. We did not measure local renal tissue cytokine levels, so the direct renal origin of these circulating signals is unproven and warrants further investigation. Third, observational associations cannot establish causality. Unmeasured variables, such as perioperative hemodynamics, nephrotoxin exposure, or subtle differences in surgical techniques, might influence both inflammation and AKI, also acting as confounders. We attempted to adjust some baseline differences, including age, diabetes and pre-operative creatinine, but unaccounted confounding is possible. The logistic models, while illustrative, were exploratory and not internally or externally validated for predictive performance. Finally, our definition of obesity (BMI ≥32) and grouping, though based on prior data, may not relate to other studies of AKI in obesity.

Our study was prospectively designed with serial sampling at multiple time points, enabling us to capture dynamic changes in biomarkers from before surgery through 4 days after surgery. Longitudinal profiling is crucial in AKI, given its rapid onset. We stratified the analysis by obesity status, addressing an often-neglected aspect of personalised risk assessment. This revealed novel differences in potential biomarker panels that could be used for diagnosis, prognosis or intervention, but would be obscured in a mixed population. The use of multivariable logistic models allowed us to identify biomarkers independently associated with AKI while controlling for key covariates. Moreover, our identified panels covered a broad array of immune pathways (chemokines, cytokines, complement, and protease inhibitors), providing a more comprehensive view of the inflammatory network and defensive counteraction in AKI post cardiac surgery.

## Data Availability

All data produced in the present study are available upon reasonable request to the authors

## Authors’ contributions

Individual contributions to the study were as follows: BY and MJW designed the study, MJW and NB wrote the manuscript with critical input from BY. GJM, LJ-D, TK and HA managed the conduct of the study. NB and HP performed the wet lab experiments. NS, BE-H and KT collected samples. MJW performed statistical analyses.

